# Insulin Resistance and Metabolic Comorbidities in Alzheimer’s Disease

**DOI:** 10.1101/2021.04.23.21255980

**Authors:** Martina Johansson

**Affiliations:** Department of Health, Medicine & Caring Sciences, Linköping University, 58185 Linköping, Sweden

## Abstract

Alzheimer’s disease affects 50 million people worldwide and just like any modern lifestyle disease, it is steadily increasing. Unfortunately, a cure has not yet been found, despite decades of research. The pathophysiology of Alzheimer’s disease is extraordinarily complex and involves many different factors. Despite it being a protein misfolding disease, a lot of evidence currently point to insulin resistance, impaired glucose metabolism and comorbidities with other metabolic disorders such as obesity, elevated blood lipids, and non-alcoholic fatty liver disease, as a potential root cause. Genetics aside, it seems like poor metabolic functioning affects the brain and central nervous system a great deal: affecting mood, behaviour, and cognitive performance. Even mood disorders are interlinked with metabolic disorders and increase the risk for Alzheimer’s disease later in life. This is a summary of objectively chosen original research articles, published between 2011-2021 and with insulin resistance as the main objective for cognitive impairment and Alzheimer’s disease.

## Introduction

Alzheimer’s disease (AD) is a metabolic disease mainly targeting the area of hippocampus, mostly responsible for short-term memory and spatial awareness.^1^ It has also been identified as a protein misfolding disease were amyloid beta (Aβ) and tau proteins are abnormally folded and accumulated as plaques and tangles. Amyloid beta precursor protein (APP) is concentrated in the synapses of neurons and regulate synapse information, neural plasticity, and antimicrobial activity. Aβ is a peptide formed from APP with several neurochemical functions but if it is not cleared it has neurotoxic effects, possibly contributing to cognitive impairment. Tau proteins also have specific functions like facilitating cellular signalling, neuronal development, neuroprotection, and apoptosis.^2^

How does this relate to metabolism? One reason for protein misfolding is disrupted phosphorylation. Phosphorylation is an important mechanism in cellular energy metabolism where proteins can be altered after they are formed. When tau and beta Aβ protein plaques are found in excess in the brain, they are not correctly folded but have instead been subject to hyperphosphorylation. This is a metabolic phenomenon related to impaired glucose metabolism and insulin response^4^. In addition, both Aβ and tau are involved in brain insulin signalling, and Aβ and insulin are substrates for the same insulin degrading enzyme. With Aβ build up and plaque formation in the brain, there is a reduction of both insulin receptor autophosphorylation as well as a reduction of insulin binding to its receptor.^3^

Several cellular and molecular mechanisms observed in AD are connected to poor insulin response and inflammation.^5^ However, inflammation and insulin resistance are closely related as elevated levels of pro-inflammatory cytokines are known to cause insulin resistance (IR). Several inflammatory cytokines, especially interleukin 1β (IL-1β) and tumor necrosis factor alpha (TNF-α), are anti-insulinemic and disrupt insulin receptor phosphorylation as well as blocking its substrates.

Patients with type 1 diabetes (T1D) that are not insulin resistant; it is an autoimmune disorder that causes impaired insulin production. Unlike patients with T2DM that have high blood sugar and high insulin levels because of insulin resistance, patients with T1D need insulin therapy to keep blood sugar levels within a target range. Both T1D and T2DM have increased levels of inflammatory biomarkers and increased risk of AD.^6^ However, the risk increases with elevated HbA1c.^7^

Patients with AD have higher levels of inflammatory biomarkers and lower levels of insulin in their cerebrospinal fluid (CSF). This indicates brain insulin resistance and is causing memory impairment by energy-starvation of the brain. It also contributes to Aβ and tau-protein build-up.^8^

Understanding metabolism in general and neurometabolism in particular, is of great importance in understanding the complex pathophysiology of AD.

## Materials and methods

Search was performed on PubMed and mesh terms included *insulin resistance, type 2 diabetes, type 1 diabetes, Alzheimer’s disease, cognitive impairment, glucose metabolism* and *neurometabolism*. The search was focused on peer-reviewed original research articles written in English, published during a 10-year period from 2011 to 2021. Articles in the form of case studies, editorials, essays, review- and meta analyses as well as articles that were not peer reviewed or published in the English language were excluded. Research articles focused on other pathophysiological phenomena than IR was also excluded. See figure 1 for screening process.

**Fig. 1.**
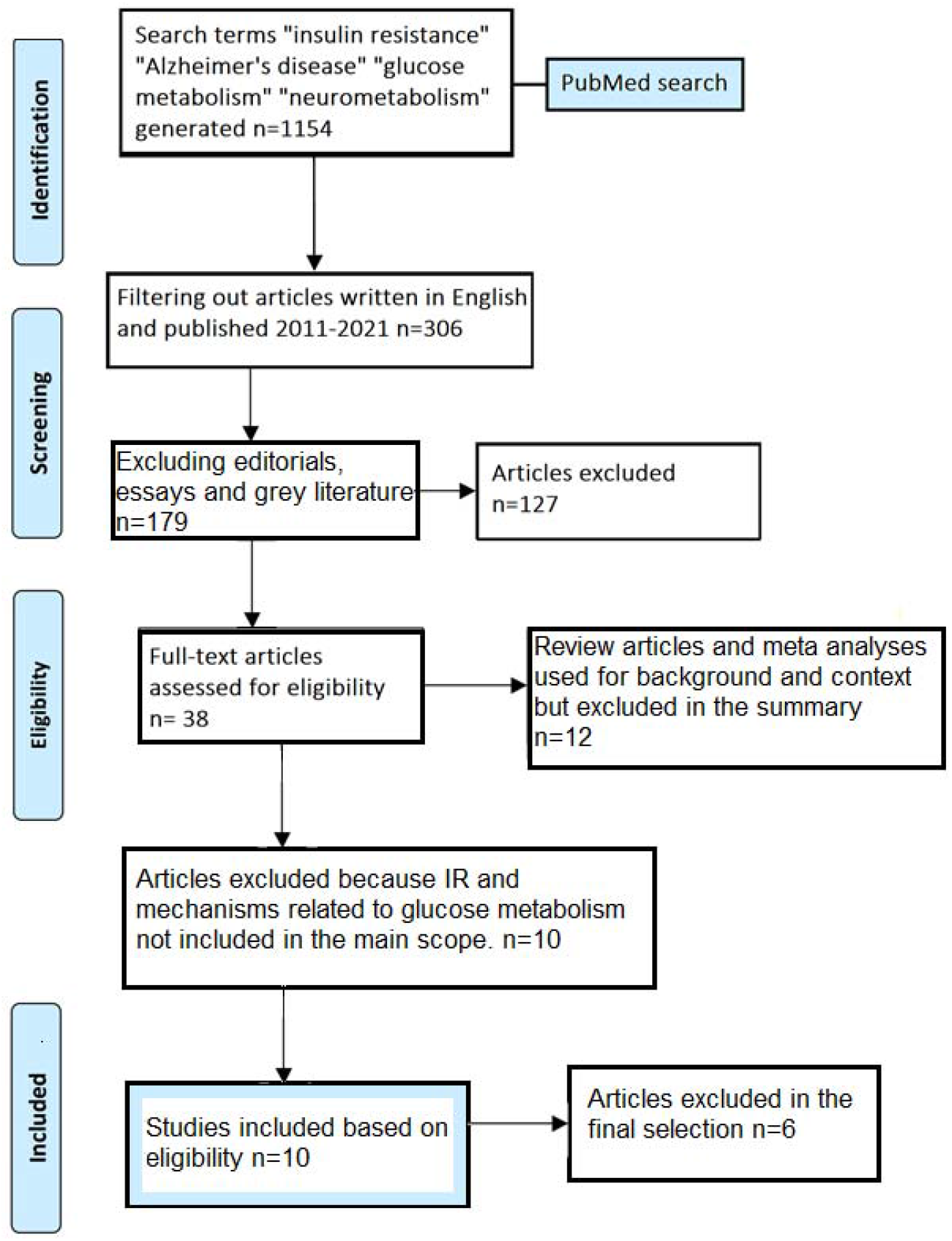
Screening process.

### Depression, insulin resistance and cognitive impairment

It is not only IR and impaired glucose metabolism that increase the risk of AD but also oxidative stress, head trauma and major depression disorder (MDD). Oxidative stress is an effect of both hyperglycaemia and inflammation and both depression and cognitive impairment (CI) is associated with higher values on the quantified homeostatic model (HOMA) used for assessing IR.^9^

The link between IR and CI goes through the hypothalamic-pituitary-adrenal (HPA) axis as its stress-hormones (cortisol) affects blood glucose levels and insulin sensitivity. Chronic mental (and physical) stress results in chronically elevated cortisol levels which prepares the body for a crisis by lowering the metabolic rate, decrease insulin sensitivity and store fat in the midsection. Complex problem solving and executive functions are associated with the prefrontal cortex and high stress, anxiety and depressive symptoms are associated with less activation in this area of the brain. Thus, mood disorders impair cognition.^10^

A cross-sectional study on 45 men and women aged 19-71, with a history of MDD but without T1D, T2D or any other documented physical or mental health problem, performed glucose- and advanced cognitive testing to see if higher steady-state plasma glucose (SSPG) was predictive of worse cognitive performance. It was not, but when the subjects were dichotomized into younger (<45 years) and older (≥45 years), SSPG was associated with poorer cognitive performance and cognitive flexibility in the younger group but not the older.^11^ One hypothesis is that metabolic syndrome has more severe consequences in younger adults, as it might imply a more fragile biochemistry. A cohort investigation over a 3–5-year period surveying random samples of patients with T2DM in a registry with 19239 participants, 30-75 years of age, showed a common comorbidity of depression and diabetes and a 100% risk increase of dementia if diagnosed with both depression and T2DM. Compared to diabetes alone, statistically at least, mood problems in patients with IR substantially increase risk for CI and AD.^12^ The correlation between HPA-axis dysregulation in both cognitive impairment and metabolic syndrome is well documented.^13^

### Liver enzymes and Alzheimer’s disease

Not just IR but several other aspects of metabolic dysfunction links to diseases in CNS. Non-alcoholic fatty liver disease (NAFLD) that almost always co-occurs with IR has a common comorbidity with CI, depression, and anxiety. A population-based study on liver enzymes and early AD progression was performed by measuring relevant metabolic factors and cognitive performance. Covariates like age, sex, BMI, education level and APOE-ε4 status (important genetic risk marker) was confounded. Patients were evaluated by CSF, MR, cognition tests and liver enzymes like ASAT and ALAT and increased liver biomarkers in NAFLD patients directly correlated with poor cognition and Aβ deposition in brain areas.^14^ Peripheral organs are affected by a dysfunctional glucose metabolism and if damage their circulation is affected. Kidneys and liver are involved in Aβ clearance. NAFLD inhibits the detoxification process of the liver which further promotes metabolic dysfunction.^15^

### Exercise and improved glucose metabolism improve cognition

Lifestyle interventions for obesity and metabolic disorders are equally beneficial for mental and cognitive disorders. Several studies have been performed on various types of exercise and cognition, both strength training (which is known to increase insulin sensitivity)^16^, aerobic exercise (which is known to lower blood glucose levels)^17^ and stretching (which is known to positively affect CNS)^18^. A Brazilian study compared two groups of older, insulin resistant adults with and without dementia. The participants performed submaximal exercise stress-tests with insulin growth factor 1 (IGF-1) being measured before and after the tests. IGF-1 increased significantly after exercise in the AD group while remained unaltered in the normal group. This was partially explained by IGF-1 resistance and inadequate functioning of IRS-2 insulin receptor substrate, related to low-grade inflammation that induces the synthesis of TNF-α. Increased IGF-1 has been associated with improvement of cognitive function in response to exercise. Evidence indicates a role for IGF-1 in beta-amyloid clearance and reducing hyperphosphorylation of tau in AD, thus this patient group could greatly benefit from increasing their cardiovascular fitness level.^19^

Another aspect of physical exercise is that a sedentary lifestyle fuels both IR, inflammation, and chronic stress.^20^

A study focused on cardio training and cognition was performed on 28 insulin resistant adults 57-83 years of age without AD. The participants had to perform 45–60-minute cardio session 4 times per week during a 6-month period, while the control group performed stretching. Cognitive performance on tasks of executive function including selective and divided attention, cognitive flexibility, and working memory improved and circulating levels of Aβ decreased in the aerobic group relative to controls.^21^ Despite it being a small study, there’s a growing body of evidence on exercise improving cognition by increased insulin sensitivity as well as increased CBF.

### Pharmacological solutions to IR and CI

Several studies have looked at impaired glucose metabolism from a pharmacological point of view, using both metformin, insulin and other supplements with glucose lowering effect. A randomized placebo-controlled crossover study on metformin was performed on non-diabetic patients with MCI due to AD. The patients took 2000 mg of metformin for 8 weeks followed by 8 weeks of placebo and metformin was associated with improved executive functioning as well as improved learning, memory, and attention. Lumbar puncture showed no difference in CSF, but MRI showed increased cerebral blood flow (CBF).^22^

Curcumin is a bioactive compound previously analysed in the context of reducing inflammation and oxidation. Animal studies show a reduction of inflammatory markers such as COX-2, NkB, TNF-α, IL-1 and CRP and a randomized controlled trial on patients with T2DM and AD showed promising results when analysed for circulating glycogen kinase 3β and insulin resistance, when ingesting 180 mg curcumin per day for 12 weeks. Curcumin also seemed to mitigate Aβ and tau build-up.^23^

Key factors for cognitive function seem to be to get energy, nutrients and oxygen delivered to the brain and another way to facilitate this is via intranasal insulin. Both regular insulin and long-lasting analogues has been used in trials and in one study, 60 adults diagnosed with MCI or mild to moderate AD received placebo, 20 IU or 40 IU of insulin for 21 days administered nasally. APOE-ε4 is the single most important genetic risk factor for AD, and patients with this genetic variation had higher insulin levels at baseline but also responded better to insulin treatment. Non-carriers experienced worsening of AD symptoms with intranasal insulin, but the highest dose (40IU) yielded improved working memory (but worse metabolic status) independent of APOE status.^24^

It seems like APOE-ε4 carriers and individuals with low overall insulin sensitivity, can benefit from insulin treatment, but the draw-back is that pro-longed exposure to insulin will worsen the metabolic health in most individual. Being metabolically robust is highly protective against cognitive decline and the difference between the APOE isoforms seem to be how they modulate glucose.

The brain relies almost exclusively on glucose for energy production which is why it is particularly sensitive to impaired glucose metabolism or IR. A study on mice with human APOE-ε2, APOE-ε3 and APOE-ε4 was conducted to compare the different brains and APOE-ε4 (but also APOE-ε3) showed markedly reduced levels of IGF-1, insulin substrate and glucose transporter 4 (GLUT-4, responsible for cellular glucose uptake) compared to APOE-ε2. APOE-ε2 is a relatively new and rare genetic variation with more robust metabolic qualities that seem to be protective against AD. This provides another strong indication that poor energy metabolism in the brain leads to cognitive impairment.^25^

### Dietary intervention for IR and AD

If energy deficit is the hallmark of AD, what other types of strategies could be beneficial to provide energy to a starving brain? A recent study from New Zealand published in *Alzheimer’s Research & Therapy*, was the first randomized crossover trial on Alzheimer patients ever made. They randomly assigned AD patients to a high fat, ketogenic diet for 12 weeks. The control group ate a healthy low-fat diet, and the ketogenic group ate 80E% fat, 15E% protein and a maximum of 5E% carbohydrates in every meal, forcing the body and brain to run on ketones instead of glucose. When completely starved of carbohydrates the body makes the small amount of glucose it needs from amino acids and ketone bodies (a by-product of the breakdown of fatty acids and β-oxidation in the liver) takes over as main energy substrate, independently of insulin. When the AD brain does not solely depend on glucose for its energy needs, cognitive function and quality of life improves.^26^ Several studies on the ketogenic diet in AD has been carried out but one drawback for this type of treatment is that it requires a high level of motivation and cooperation. Cognitive impairment undoubtedly negatively impacts these faculties, making it a lot harder to implement than a pharmacological solution. Despite this, the ketogenic diet has several benefits for the AD brain. Lowering glucose requirements is one thing, providing another fuel is another, but it also seems to lower inflammation and promote phosphorylation and stabilized synaptic function.^27^

## Discussion

In recent years strict low carb diets have been approved by American Diabetes Association (ADA) and European Association for the Study of Diabetes (EASD) as a successful treatment method for IR and T2DM.^28^ Reversing IR decreases the risk of developing AD, even with genetic factors included. However, most people have difficulties adhering to a strict diet, especially AD patients, even if doing so has a great chance of improving quality of life. This poses a serious challenge for the development of future therapies in treating AD. More information on how different foods as well as lifestyle choices affect the entire organism is needed.

HPA dysregulation, stress, depression, and anxiety is extremely common as mental health problems are on the rise. Together with an increase in metabolic syndrome and insulin resistance, it makes sense to view mental and metabolic disorders in conjunction as they might be interlinked with a suboptimal lifestyle as the common denominator. Being proactive or treating illnesses as early as possible is of utter importance, especially in younger people. Early development of various diseases including IR and mental health problems signifies a more fragile biochemistry, increasing susceptibility to CI and AD later in life.

Reliable pharmacological solutions are needed, but since the progression of effective AD medication is slow, lifestyle interventions seem to be the fastest route to prevention as also for improving the wellbeing of patients already diagnosed with AD.

## Conclusion

Several pieces of evidence have demonstrated that an unhealthy or improper lifestyle that leads to poor mental and metabolic health impact CNS function through the gut-brain axis, HPA axis and brain metabolism. The metabolic syndrome seems to worsen amyloid burden due to brain energy starvation and ineffective Aβ clearance which in turn increase neuroinflammation. Using pharmacological interventions such as intranasal insulin is a poor crutch compared to social support and effective lifestyle interventions with physical exercise, social activities, and a diet with low glycaemic load. More studies with focus on different types of lifestyle interventions and AD is clearly needed.

## Data Availability

N/A

## Declarations

### Ethics approval and consent to participate

N/A

## Consent for publication

N/A

## Availability of data and material

N/A

## Author Contributions

Martina Johansson came up with the idea of the review, performed the search and analysed the retrieved papers with the purpose of expanding knowledge in the field of neurometabolism in AD.

## Acknowledgements

N/A

## Appendix

**Table. 1.** Overview and summary of the objectively selected original research articles.

| Journal | Title | year | author |
| --- | --- | --- | --- |
| *Journal of psychiatric research* | Association between insulin resistance and cognition in patients with depressive disorders: Exploratory analyses into age-specific effects abnormalities and behaviour. | **2015** | Wroolie T et al. |
| *Archives of general psychiatry* | Association of depression with increased risk of dementia in patients with type 2 diabetes: The Diabetes and Aging Study. | **2012** | Katon W et al. |
| *JAMA network open* | Association of altered liver enzymes with Alzheimer's disease diagnosis, cognition, neuroimaging measures, and cerebrospinal fluid biomarkers. | **2019** | Nho K et al. |
| *Behavioural Brain Research* | Acute exercise increases circulating IGF-1 in Alzheimer's disease patients, but not in older adults without dementia. | **2021** | Stein A et al |
| *Journal of Alzheimer's Disease* | Aerobic exercise improves cognition for older adults with glucose intolerance, a risk factor for Alzheimer's disease. | **2011** | Baker L et al |
| *Alzheimer's disease and associated disorders* | Effects of the insulin sensitizer metformin in Alzheimer's disease: Pilot data from a randomized placebo-controlled crossover study. | **2017** | Koenig A M et al |
| *Nutrients* | Dietary Supplementation with Curcumin Reduce Circulating Levels of Glycogen Synthase Kinase-3β and Islet Amyloid Polypeptide in Adults with High Risk of Type 2 Diabetes and Alzheimer's Disease | **2020** | Thota R et al. |
| *Journal of Alzheimer's Disease* | Long-acting intranasal insulin detemir improves cognition for adults with mild cognitive impairment or early-stage Alzheimer's disease dementia. | **2015** | Claxton A et al |
| *Alzheimer's research & therapy* | Randomized crossover trial of a modified ketogenic diet in Alzheimer's disease. | **2021** | Phillips M et al. |

## Abbreviations

AD: Alzheimer’s Disease
Aβ: Amyloid beta
APP: Amyloid beta precursor protein
CBF: Cerebrospinal blood flow
CSF: Cerebrospinal spinal fluid
CI: Cognitive impairment
HPA: Hypothalamus, Pituitary, Adrenal
HOMA-IR: Homeostatic model for insulin resistance
IGF-1: Insulin growth factor 1
IR: Insulin resistance
MCI: Major cognitive impairment
MDD: Major depressive disorder
NAFLD: Non-alcoholic fatty liver disease
SSPG: Steady-state plasma glucose
T2DM: Type 2 Diabetes Mellitus
T1D: Type 1 Diabetes
TNF-α: Tumor necrosis factor alpha

